# Promoting Activity, Independence and Stability in early dementia and mild cognitive impairment (PrAISED): A randomised controlled trial

**DOI:** 10.1101/2022.12.20.22283699

**Authors:** Rowan H Harwood, Sarah E Goldberg, Andrew Brand, Veronika van Der Wardt, Vicky Booth, Claudio Di Lorito, Zoe Hoare, Jennie Hancox, Rupinder Bajwa, Clare Burgon, Louise Howe, Alison Cowley, Trevor Bramley, Annabelle Long, Juliette Lock, Rachael Tucker, Emma Adams, Rebecca O’Brien, Fiona Kearney, Katarzyna Kowalewska, Maureen Godfrey, Marianne Dunlop, Kehinde Junaid, Simon Thacker, Carol Duff, Tomas Welsh, Annette Haddon-Silver, John Gladman, Pip Logan, Kristian Pollock, Kavita Vedhara, Victoria Hood, Roshan Das Nair, Helen Smith, Rhiannon Tudor-Edwards, Ned Hartfiel, Victory Ezeofor, Robert Vickers, Martin Orrell, Tahir Masud

## Abstract

**Background:** Dementia is associated with frailty leading to increased risks of falls and hospitalisations. Interventions are required to maintain functional ability, strength and balance.

**Design:** Multi-centre parallel group randomised controlled trial, with embedded process evaluation. Procedures were adapted during the COVID-19 pandemic.

**Participants:** People with mild dementia or mild cognitive impairment (MCI), living at home, and a family member or carer.

**Objectives:** To determine the effectiveness of an exercise and functional activity therapy intervention compared to usual care.

**Intervention:** A specially-designed dementia-specific rehabilitation programme focussing on strength, balance, physical activity and performance of ADL, which was tailored, progressive, addressed risk and the psychological and learning needs of people with dementia, providing up to 50 therapy sessions over 12 months. The control group received usual care plus a falls risk assessment.

**Main outcome measure:** The primary outcome was the informant-reported Disability Assessment for Dementia (DAD) 12 months after randomisation. Secondary outcomes were: self-reported ADL, cognition, physical activity, quality of life, frailty, balance, functional mobility, fear of falling, mood, carer strain and service use (at 12 months) and falls (between months 4 and 15).

**Results:** 365 people were randomised, 183 to intervention and 182 to control. Median age of participants was 80 years (range 65-95), median Montreal Cognitive Assessment score 20/30 (range 13-26), 58% were men. Participants received a median of 31 (IQR = 22-40) therapy sessions out of a possible maximum of 50. Participants reported completing a mean 121 minutes/week of PrAISED activity outside of supervised sessions. Primary outcome data were available for 149 (intervention) and 141 (control) participants. There was no difference in DAD scores between groups: adjusted mean difference -1.3/100, 95% Confidence Interval (−5.2 to +2.6); Cohen’s d effect size -0.06 (−0.26 to +0.15); p=0.5. Upper 95% confidence intervals excluded small to moderate effects on any of the range of secondary outcome measures. Between months 4 and 15 there were 79 falls in the intervention group and 200 falls in the control group, adjusted incidence rate ratio 0.78 (0.5 to 1.3); p= 0.3.

**Conclusion:** The intensive PrAISED programme of exercise and functional activity training did not improve ADLs, physical activity, quality of life, reduce falls or improve any other secondary health status outcomes even though uptake was good. Future research should consider alternative approaches to risk reduction and ability maintenance.

**Trial registration:** ISRCTN15320670.

**Funding:** National Institute for Health and Care Research

**What is already known:**

- Dementia is associated with progressive loss of functional ability, including activities of daily living and mobility, and a high risk of falls
- Exercise programmes and rehabilitation therapies may improve ability, or slow the rate of decline, but evidence from trials and systematic reviews is equivocal

**What this study tells us:**

- We developed an intensive dementia-specific exercise and functional activity rehabilitation programme, lasting 12 months, taking account of motivation, learning needs and context, in particular the need to engage carers, and evaluated it in a randomised controlled trial
- The programme was very well received by participants and therapists, but had no effect on activities of daily living, physical activity, quality of life, falls, cognition or any other health status outcome
- We are unlikely to be able to change rate of loss of ability in dementia through exercise or functionally orientated rehabilitation therapy.
- We need different ways of defining wellbeing after a dementia diagnosis.

## INTRODUCTION

Dementia causes progressive loss of ability in activities of daily living (ADL) and physical activity. Multiple mechanisms lead to functional loss, including cognitive and neurological decline, co-morbidities, acute illness, injuries, delirium, inactivity, deconditioning and restriction of opportunities, especially if the person suffers stigma or family members are concerned for safety. Dementia (and a possible precursor state, mild cognitive impairment, MCI) confers an increased risk of crises, including acute physical illness and a twofold increased risk of falls. Sixty to 80% of people living with dementia fall each year [1-5].

Dementia prevalence increases exponentially with age, affecting 20% of 80-year-olds. A similar number again have MCI [6,7]. Prevalence in the population will double in the next 30 years [8]. Dementia is one of the main drivers of dependency (the need for help from other people). It creates high levels of demand on health and social care services, families and other informal carers [9]. The English National Dementia Strategy emphasised the importance of early diagnosis and the goal of living well with dementia [10]. Reasons to diagnose dementia include access to cognitive enhancing drugs and cognitive stimulation therapy, however their effects are small [11,12]. Commentators have highlighted a relative lack of available therapeutic interventions [13]. Exercise has been proposed as a way of preventing or slowing the progression of dementia [14], but in trials, has been shown to have little effect on global cognition [15-18]. Some evidence suggests that exercise might slow the decline in ADL performance or prevent falls [15,18,19].

We hypothesised that exercise-based, functionally-directed rehabilitation would improve physical reserve, promote safe performance of activities, reduce falls, enhance recovery from intercurrent illness or injury, and hence improve ADL. We undertook a programme of research to develop and evaluate this approach [20]. We developed a dementia-specific therapy intervention, called Promoting Activity, Independence and Stability in Early Dementia and mild cognitive impairment (PrAISED) [21]. The target population was those with relatively mild impairment who might retain capacity to learn, change behaviour and develop new routines. We conducted a 3-arm feasibility trial with 60 participants, comparing the PrAISED intervention delivered with 12 months supervision, a reduced schedule of 3 months supervision, and a control condition, in which we demonstrated that intervention delivery and research were feasible and found benefits in balance and mobility outcomes, but most participants required ongoing supervision to sustain adherence [21-23]. The aim of this definitive trial was to determine the effectiveness of the PrAISED intervention on ADL, falls, physical activity and quality of life, among other outcomes.

## METHODS

### Study design

We performed a multi-centre, individually 1:1 randomised, stratified, pragmatic, parallel group, randomised controlled trial [24]. The trial was impacted by the COVID-19 pandemic. Recruitment was paused and delivery adaptations instituted between March and September 2020.

### Study setting

Participants were recruited by trained researchers from five sites in England via secondary care memory clinics (dementia diagnostic services), general practice registers, dementia support groups, and the NIHR *Join Dementia Research* register. The intervention was delivered in participants’ homes and local communities.

### Participants

Patient participants were aged over 65 years, had a diagnosis of MCI or dementia, a Montreal Cognitive Assessment (MoCA) score of 13-25 (out of 30), a family member or carer who knew the participant and had a minimum of 1-hour weekly contact in person or by telephone/internet, and was willing to act as an informant. Participants had to be able to walk without human help, communicate in English, have adequate sight, hearing, and dexterity to complete neuropsychological tests, mental capacity to give consent as assessed by a study researcher, and consented to participate. Carers participated in their own right, and separate consent was taken.

Exclusion criteria included a diagnosis of Dementia with Lewy Bodies, a comorbidity preventing participation (such as severe breathlessness, pain or severe neurological disorder), anticipated unavailability over the next year (e.g., relocation, prolonged holiday) or life expectancy less than a year.

### Baseline data

The study dataset included multiple health status measures, as appropriate for a complex intervention trial [25-30]. The rationale was to measure a range of credible predictor, mediator, intermediate and distal health status outcomes, including ADL, balance, mobility, frailty, executive function, mood, carer strain and quality of life.

Baseline data comprised demographics, medications, medical and falls history, ADL (Disability Assessment for Dementia, DAD [31]; Nottingham Extended ADL scale, NEADL [32]); cognition (MoCA [33]); animal-naming verbal fluency; Cambridge Neuropsychological Test Automated Battery, CANTAB [34]); mood (Hospital Anxiety and Depression Scale, HADS [35]); Apathy Evaluation Scale [36]; physical activity (Longitudinal Aging Study of Amsterdam Physical Activity Questionnaire, LAPAQ [37,38]); step count (Misfit Shine accelerometer); self and proxy-assessed quality of life (DEMQOL [39], DEMQOL-U [40], EuroqoL EQ-5D-3L [41]); fear of falling (short Falls Efficacy Scale-International, FES-I [42]); frailty (SHARE index [43]); balance (Berg Balance Scale [44]); mobility and ability in divided attention (Timed Up and Go, TUG, dual-task TUG [45]); hand grip strength (Camry EH101 Electronic Hand Dynamometer); health and social care resource use for patient and carer (Client Service Receipt Inventory, CSRI [46]); carer strain (Caregiver Strain Index, CSI [47]); carer health-related quality of life (EQ-5D-5L). Verbal fluency, apathy and CANTAB measures were intended as markers of executive function, which is associated with risk of falls, and has been reported to improve with exercise interventions [48-51].

### Intervention

The intervention was delivered by National Health Service (NHS) and other local healthcare providers, according to a manual [52](TIDieR checklist, Appendix 1). Clinicians were trained by centrally-based research therapists, including a two-day initial course, a mid-point refresher conference, and weekly teleconferences to discuss problems and difficult scenarios.

The development and content of the PrAISED intervention have been published [21,52,53]. Participants in the intervention arm received an individually tailored programme comprising physical exercises (i.e., progressive strength, balance, and dual task training [54-58]; functional activities (i.e., ADL with an element of physical activity, such as going out shopping)[59, 60]; inclusion in community life (e.g., through signposting physical exercise classes and exercise facilities); risk enablement (positive risk-taking); [61] and environmental assessment (accessibility and safety issues at home). Participants received up to 50 home therapy sessions over 12 months from a multidisciplinary team comprising physiotherapists, occupational therapists and rehabilitation support workers (associate practitioners). Sessions were intended to teach and supervise exercise and functional activities, monitor progress and adjust and progress the programme. Delivery used a specifically developed behaviour change model [23, 62-67]. The intervention was tailored to individual abilities, co-morbidities, interests, and goals using a stratification tool to determine the frequency of intervention to enable participants to sustain the programme [52]. Participants were encouraged to perform their programme for a minimum of three hours per week based on previous research findings for improvements in falls and executive function [48-50, 68, 69]. Family members or carers were encouraged to support or participate where possible. The amount of supervision was ‘tapered’ (became progressively less frequent over time) to encourage habit formation and promotion of self-directed exercise and activity between supervised sessions and after the programme had finished. Therapy sessions were intended to be delivered in-person.

The control intervention consisted of a falls prevention assessment and advice and was modelled on usual falls prevention care. This consisted of an initial therapy visit for assessment and up to two further visits by a study therapist to review actions, give advice, and refer on to their general practitioner (GP) or local services, if needed. These visits lasted up to 90 minutes. The control participants were seen by the same therapists who delivered the active intervention.

Both study groups were assessed using the Guide to Action falls risk factor assessment tool [70]. Advice was offered based on the findings including further clinical assessment, assessment for equipment, and medication review by their GP, if necessary. Any non-study intervention was permitted in both study arms including cognitive stimulation therapy, use of acetylcholine inhibitor or memantine drugs, or referrals to mental health, medical, rehabilitation or falls prevention services.

### Outcome evaluation

Each participant took part for 15 months. A brief postal follow up, with telephone support if needed, was undertaken with a carer after 6 months. The main follow up was completed at 12 months (+/-4 weeks) when participating dyads were visited at home by two researchers to collect outcome data by interviewing the participant and carer separately (table 1). This follow up was undertaken remotely via telephone or video-calls during the COVID-19 lockdown.

**Table 1:**
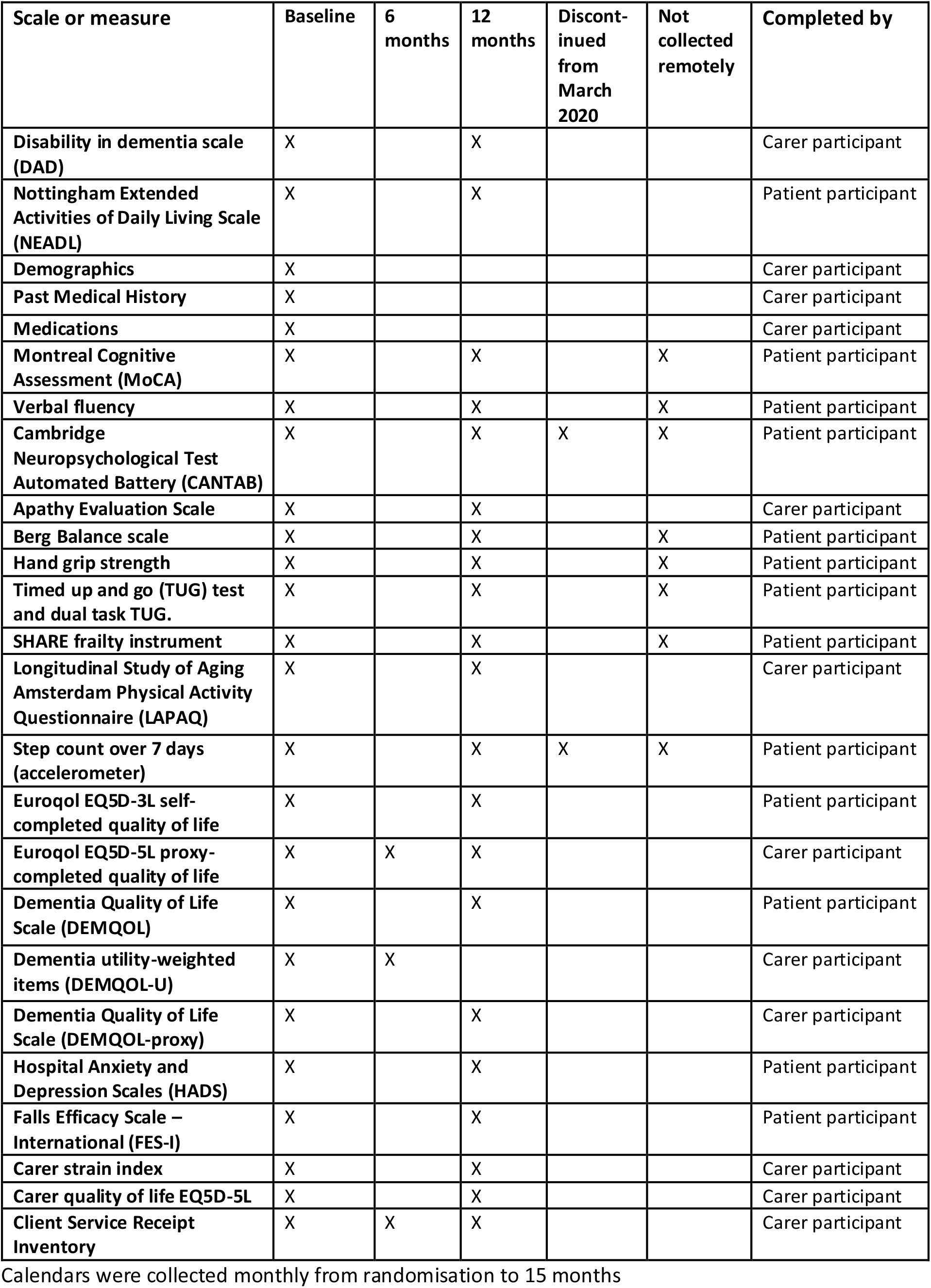
Data collection time points.

Falls, PrAISED activities done independently, service use and hospitalisations were ascertained using monthly self-completed calendars between months 0 and 15 with telephone prompts and support by researchers not involved in delivery of the intervention, where necessary. Injurious falls were adjudicated by two clinicians based on details provided on calendars.

### Study outcomes

At 6 months, quality of life (EQ-5D-3L, DEMQOL-U) and service use (short CSRI) were collected.

The primary outcome was carer-informant-rated disability in ADLs, measured by the DAD, after 12 months. Secondary outcomes at 12 months were the self-reported NEADL; falls, rate of falling and injurious falls; cognition (MoCA, verbal fluency, apathy evaluation scale and CANTAB); quality of life (DEMQOL, EQ5D); activity (LAPAQ, accelerometers); frailty (SHARE Index); Berg Balance Scale, functional mobility (single and dual task TUG test), hand grip strength; fear of falling (FES-I); mood (HADS); carer strain (CSI) and carer quality of life (table 1).

### Harm and Adverse events

Adverse events were classified as serious (death, life-threatening events, hospitalisation, significant incapacity), or potentially related to the intervention or to PrAISED exercises undertaken independently. Adverse events were ascertained by participants or their carer reporting them to the study or intervention delivery teams or through monthly calendars. The intervention group had more exposure to study therapists, and consequently more opportunity to report adverse events, resulting in an ascertainment (information) bias. To compare the safety of the intervention we considered deaths, hospital admissions and falls to be core adverse events. To investigate the possibility of an early falls hazard associated with increased activity, we analysed falls in the first three months separately.

### Impact of the Covid-19 pandemic

Following UK Government guidance for strict social distancing and the requirement to remain at home, all non-essential face-to-face contact ceased on 17th March 2020. At this time, 64 participants had completed the study, 187 were in process and 27 had been recruited but had not commenced therapy. A series of mitigating measures was undertaken and PrAISED therapists were provided with guidance (Appendix 2). A protocol amendment to adapt trial procedures was approved, including delivery of the intervention via telephone or videocall [71]. Follow up assessments were conducted remotely, meaning we could not complete measures requiring physical contact. We removed some outcome measures to reduce burden on participants (Table 1).

Participants within 6 weeks of the end of their programme had their final assessment brought forward. An additional interim outcome data collection point was introduced for all other remaining participants, in case no further trial activity became possible. Recruitment and in-person therapy and data collection recommenced after 1^st^ September 2020, if participants were willing, using personal protective equipment, and excluding assessments that required close personal contact or sharing of equipment (including CANTAB cognitive measures and accelerometers). Some remote assessment continued after this time if requested by the participant.

### Sample size

An initial calculation based on parameters from published literature suggested a sample size of 368 participants (184 per group), with a 23% attrition rate, had 80% power to detect changes in disability outcome (DAD), with an effect size of 0.5 (11 points on a baseline of 70, standard deviation 22) [19, 72]. A minimum clinically important difference has not been defined for the DAD, but a natural history study in Alzheimer’s disease suggested the loss of about one point per month over 12 months [73]. Following the feasibility study, a recalculation suggested that a sample size of 248 was sufficient. The original sample size was maintained, with the agreement of the data monitoring and steering committees, in the light of uncertainties in estimates.

In the event, with the COVID-19 pandemic, this proved prescient. Recalculation of sample sizes in July 2020, prior to restarting the trial, under a range of feasible impacts on intervention effect size, primary outcome standard deviation and withdrawal rates, suggested that the sample size of 368 had adequate power to answer the research question.

### Randomisation

Randomisation was performed after baseline assessment and consent. A secure internet-based system using a dynamic, adaptive allocation algorithm, accessed by a secure web portal held at the North Wales Organisation for Randomised Trials in Health Clinical Trials Unit, Bangor University was used to randomise individuals, 1:1, minimised by site, presence of a co-resident, and history of previous falls [74]. The randomisation system was maintained by a statistician independent of the analysis and research teams. Allocation was later emailed to the intervention delivery teams, who arranged for the first clinical assessment.

### Blinding

Blinding of participants and therapists was not possible due to the nature of the intervention. Researchers collecting outcome data were not blinded as participants almost always inadvertently unblinded the researcher during the feasibility study [22]. Analysis was blinded.

### Statistical Methods

For the primary outcome of difference in DAD score between groups, an analysis of covariance (ANCOVA) was conducted using stratification variables (site, co-resident carer and history of falls) and baseline DAD score as covariates. For secondary outcome measures, ANCOVAs were conducted, using the stratification and respective baseline measures as covariates. All analyses were conducted on an intention-to-treat basis. Multiple imputation using chained equations (MICE) was used in the main analysis when appropriate. Effect sizes were standardised as Cohen’s d [75]. Minimum clinically important differences on scores were identified, where available. Adjusted mean differences, effect size estimates, 95% confidence intervals (CI) and p-values were reported for all analyses. A range of sensitivity analyses was performed, including a complete case analysis. Falls were analysed as the proportions of participants falling, the incidence rate ratio using a negative binomial regression, and time to first fall using a Cox proportional hazards regression. We anticipated that any impact of the PrAISED intervention on falls would not be immediate, so our pre-defined efficacy outcome was rate of falling between months 4 and 15. The statistical analysis plan is available [76].

### Process Evaluation

We undertook a process evaluation following Medical Research Council (MRC) guidelines [77-80]. We investigated reach, dose, fidelity and adaptations of training and intervention delivery. We recorded details of each session delivered, exercise undertaken independently via monthly calendars, and fidelity of delivery from analysis of a sample of 14 video-recorded sessions, in which evidence was sought of 14 key principles of the PrAISED intervention [52]. We conducted qualitative interviews with a sample of participants with dementia, carers and therapists to investigate how the intervention was received, as well as barriers and facilitators to participation.

### Ethics and study monitoring

The study was approved by the Bradford-Leeds Research Ethics Committee (REC number 18/YH/0059). The trial was monitored by an independent steering committee and a data monitoring committee [81].

### Patient and public involvement

Public and patient involvement (PPI) was integrated into every stage of the research cycle with the aim that the intervention had relevance and the research processes were acceptable to people with mild dementia and their carers. One of our co-investigators was a carer. PPI representatives were members of the programme management group and the trial steering committee. They worked in collaboration with the research team to develop the funding application and intervention, co-designed patient facing materials, participated in research interviews [82] and interpreting results.

## RESULTS

### Participant Flow

From 8th October 2018 to 23rd June 2022, 1540 potential patient participants were pre-screened, of whom 319 were ineligible and 746 did not wish to take part. Of 475 screened, 110 were not randomised: 61 were ineligible, 18 withdrew, 31 were lost (figure 1). 365 patient and 365 carer participants were randomised (84, 23% Bath; 79, 22% Derby; 60, 16% Lincoln; 23, 6% Oxford; 119, 33% Nottingham). Three protocol deviations involved failure to adjust baseline MoCA scores of 25/30 for duration of education.

**Figure 1:**
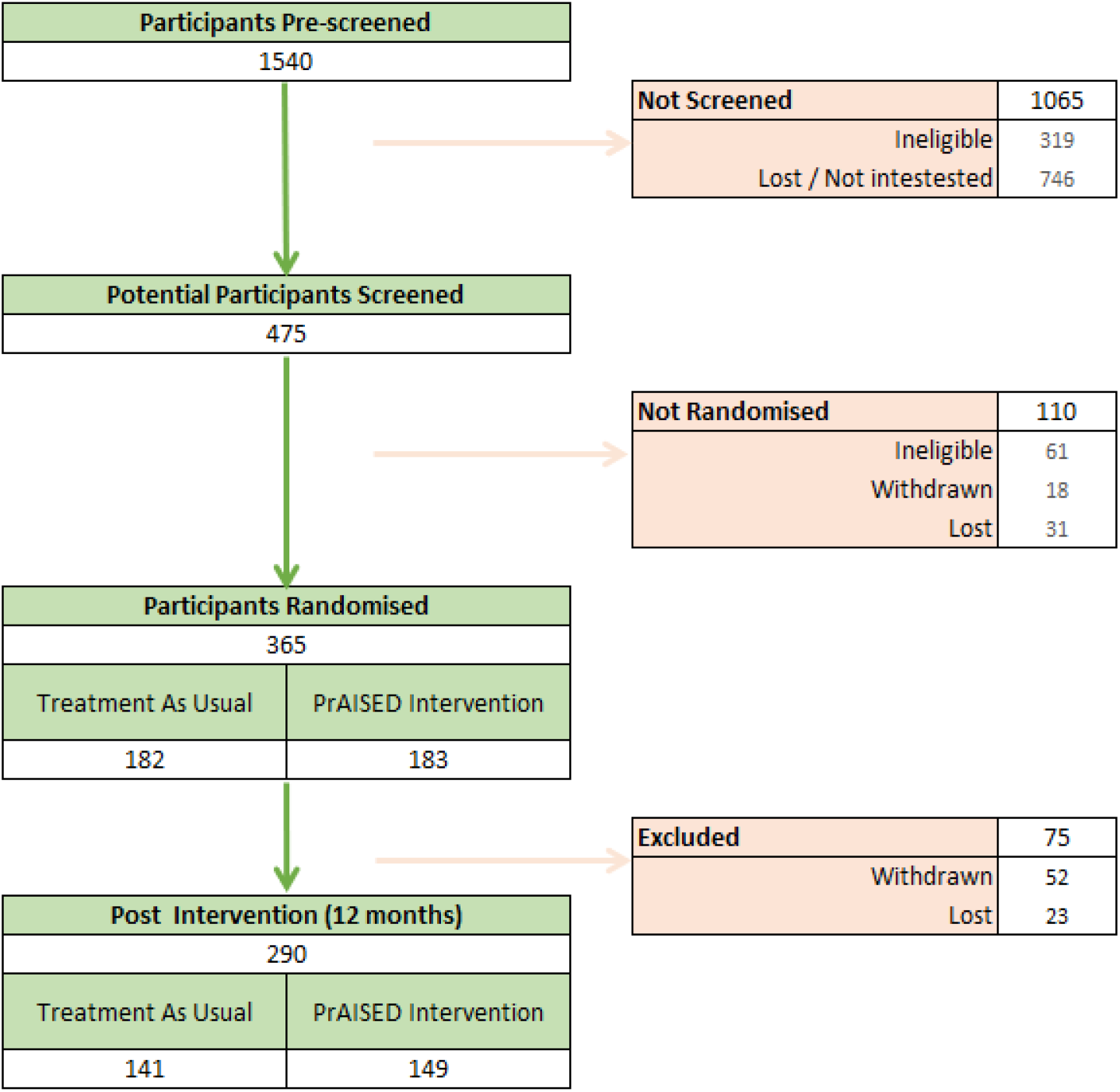
Consort Participant Flow Diagram.

### Numbers analysed

Seventy-five participants (21%) did not complete 12-month follow up: 52 (14%) withdrew, 23 (6%) were lost. There was no difference in withdrawals between groups (26 vs 26, Fischer’s Exact Test p=0.9). Available details for participants who withdrew were assessed by a blinded panel for whether they had ‘meaningful’ health-related outcomes, that is, had died, been admitted to a care home or withdrew due to deteriorating health. Twenty-seven withdrawals were meaningful, with no difference between groups (14 vs 13, p=1.0).

### Baseline Data

Baseline characteristics were similar between groups, including those who withdrew (table 2). Patient participants had a median age of 80 years (range 65 to 95), 58% male, 98% white ethnicity, 68% married and 76% co-resident with their carer. A third (31%) completed a college or university degree. 19% had MCI, 39% Alzheimer’s dementia, 19% vascular dementia, 16% mixed dementia, 7% other or unknown. Carer participants’ median age was 70 years (range 20 to 94), mostly spouses (65%) and predominantly female (73%). A third (34%) of carers had a limiting long-term health condition.

**Table 2:** Baseline variables split by allocation group.

|  | **Control**<br>**N= 182** | **Intervention**<br>**N= 183** | **Withdrew**<br>**n=52** |
| --- | --- | --- | --- |
| **PATIENT PARTICIPANTS** |  |  |  |
| **Age** median (IQR)/years | 81(75-84) | 80 (75-85) | 80 (77-85) |
| **Gender** Female n (%) | 73 (40%) | 82 (45%) | 23 (44%) |
| **Ethnic Group** White n (%) | 179 (98%) | 179 (98%) | 52 (100%) |
| **Married** n (%) | 123 (68%) | 124 (68%) | 34 (66%) |
| **Living Alone** n (%) | 46 (25%) | 43 (24%) | 17 (33%) |
| **Montreal Cognitive Assessment (MoCA)** mean/30 (SD) | 19.8 (3.1) | 20.0 (3.2) | 19.9 (3.2) |
| **Higher education** | 54 (30%) | 59 (32%) | 12 (23%) |
| **Diagnosed dementia** | 142 (78%) | 151 (83%) | 44 (85%) |
| **Comorbidity count/23** Mean (SD) | 4.0 (2.0) | 3.9 (1.8) | 3.9 (1.9) |
| **Number of drugs** Mean (SD) | 6.1 (3.5) | 6.1 (3.2) | 6.1 (3.4) |
| **Previous fall** | 95 (52%) | 93 (53%) | 26 (50%) |
| **CARER PARTICIPANTS** |  |  |  |
| **Husband/wife/partner** | 119 (65%) | 117 (64%) | 32 (62%) |
| **Son/daughter** | 50 (28%) | 55 (30%) | 16 (31%) |
| **Other** | 13 (7%) | 11 (6%) | 4 (8%) |
| **Carer Gender** Female | 134 (74%) | 131 (72%) | 38 (73%) |
| **Carer co-resident** | 136 (75%) | 140 (76%) | 38 (73%) |
| **Carer long term medical condition** | 63 (35%) | 61 (33%) | 20 (39%) |
| **Carer Age** (median, IQR)/years | 70 (58-77) | 70 (58-78) | 69 (58-78) |

IQR Interquartile Range, SD standard deviation.

### Adherence and fidelity

The intervention was delivered largely as intended and participants engaged well, despite the disruption of the COVID-19 pandemic. Participants in the intervention group took part in a median of 31 therapy sessions (IQR=22 to 40). Participants in the control group took part in a median of one session (IQR=1 to 1). The mean length of sessions was 71 minutes (SD=30; range: 5 to 220). Two-thirds were delivered face-to-face (n=1,357; 68%). Fidelity of therapy delivery was 70% against PrAISED core principles rated from video-recordings. 4,040/4,863 (83%) expected calendars were returned from intervention participants. The intervention group participants recorded a mean of 482 minutes of PrAISED exercise per month (standard deviation=705; range: 0 to 5310; 121 minutes/week) (Appendix 2).

### Outcomes

There was no difference between intervention and control on the primary ADL outcome, DAD: adjusted mean difference -1.3 (95% CI -5.2 to +2.6); effect size (d) -0.06, 95% CI -0.26 to 0.15, p=0.5, or on most secondary outcome measurements, including balance, functional mobility, physical activity or quality of life (tables 3 and 4). Upper 95% confidence intervals excluded small to moderate beneficial treatment effects. There were statistically significant small differences, in favour of the control group, on the dual task Timed Up and Go test (d=0.48, 95% CI 0.12 to 0.83, p=0.01) and (self-reported) DEMQOL (d=-0.26, 95% CI -0.47 to -0.06, p=0.01), but not DEMQOL proxy or Euroqol EQ5D quality of life measures.

**Table 3:** Unadjusted scores on outcome measures according to randomisation group.

| Measure | Group | n | Baseline Mean (SD) | Follow Up Mean (SD) | Difference | Interpretation |
| --- | --- | --- | --- | --- | --- | --- |
| **ADL DAD /100**<br>Higher is superior | Control | 125 | 77.8 (20.8) | 66.4 (24.5) | -11.4 | Deterioration |
|  | Intervention | 133 | 77.6 (20.1) | 64.2 (25.7) | -13.4 | Deterioration |
| **NEADL Score/22**<br>Higher is superior | Control | 124 | 16.8 (3.9) | 14.1 (4.8) | -2.7 | Deterioration |
|  | Intervention | 129 | 16.2 (4.2) | 13.9 (4.3) | -2.2 | Deterioration |
| **Physical Activity (LAPAQ)**<br>Higher is superior | Control | 118 | 1483 (1608) | 1293 (1430) | -189 | Deterioration |
|  | Intervention | 130 | 1395 (1230) | 1037 (1224) | -358 | Deterioration |
| **Accelerometer-Total Steps in 7 days**<br>Higher is superior | Control | 22 | 21412 (20112) | 21694 (17308) | 282 | Improvement |
|  | Intervention | 21 | 24410 (21081) | 20584 (15226) | -3826 | Deterioration |
| **Berg Balance Scale/56**<br>Higher is superior | Control | 58 | 50.3 (5.5) | 46.7 (10.6) | -3.6 | Deterioration |
|  | Intervention | 66 | 46.8 (9.6) | 46.3 (9.2) | -0.5 | Deterioration |
| **Timed Up and Go /seconds**<br>Higher is inferior | Control | 69 | 13.9 (6.7) | 14.0 (7.0) | +0.1 | Deterioration |
|  | Intervention | 69 | 13.7 (4.5) | 16.6 (12.6) | +3.0 | Deterioration |
| **Dual Task Timed Up and Go/seconds**<br>Higher is inferior | Control | 62 | 18.4 (8.3) | 20.8 (9.9) | +2.3 | Deterioration |
|  | Intervention | 64 | 19.7 (11.7) | 28.1 (20.0) | +8.4 | Deterioration |
| **DEMQOL/112**<br>Higher is superior;<br>MCID 6 | Control | 136 | 90.9 (11.4) | 88.9 (14.9) | -2.0 | Deterioration |
|  | Intervention | 140 | 89.2 (12.9) | 83.7 (15.2) | -5.5 | Deterioration |
| **DEMQOL Proxy/124**<br>Higher is superior;<br>MCID 6 | Control | 135 | 95.6 (12.9) | 90.7 (15.1) | -4.9 | Deterioration |
|  | Intervention | 145 | 92.1 (13.3) | 90.6 (13.3) | -1.5 | Deterioration |
| **DEMQOL-U (6 months)**<br>Higher is superior | Control | 149 | 0.69 (0.1) | 0.72 (0.13) | +0.03 | Improvement |
|  | Intervention | 141 | 0.72 (0.1) | 0.72 (0.13) | 0 | Deterioration |
| **Self-reported Quality of life EQ-5D-3L/1.0**<br>Higher is superior | Control | 135 | 0.82 (0.18) | 0.75 (0.25) | -0.07 | Deterioration |
|  | Intervention | 138 | 0.81 (0.18) | 0.75 (0.24) | -0.06 | Deterioration |
| **Proxy Quality of life EQ-5D-5L/1.0**<br>Higher is superior | Control | 130 | 0.80 (0.18) | 0.71 (0.23) | -0.09 | Deterioration |
|  | Intervention | 143 | 0.79 (0.17) | 0.73 (0.19) | -0.07 | Deterioration |
| **Montreal Cognitive Assessment /30**<br>Higher is superior | Control | 77 | 20.0 (3.2) | 17.5 (4.6) | -2.5 | Deterioration |
|  | Intervention | 75 | 20.1 (3.5) | 17.3 (5.2) | -2.8 | Deterioration |
| **Verbal fluency/words**<br>Higher is superior | Control | 79 | 12.3 (4.7) | 10.8 (4.8) | -1.5 | Deterioration |
|  | Intervention | 76 | 12.0 (4.6) | 10.0 (4.4) | -1.9 | Deterioration |
| **Apathy Evaluation Scale / 72**<br>Higher is inferior | Control | 121 | 40.3 (12.0) | 44.6 (12.0) | +4.3 | Deterioration |
|  | Intervention | 134 | 42.4 (12.4) | 46.3 (13.0) | +3.9 | Deterioration |
| **Falls Efficacy Scale – International/28**<br>Higher is inferior | Control | 134 | 10.3 (4.1) | 10.9 (4.8) | + 0.5 | Deterioration |
|  | Intervention | 138 | 10.4 (3.9) | 11.0 (4.5) | +0.6 | Deterioration |
| **HADS Anxiety/21**<br>Higher is inferior | Control | 133 | 3.7 (2.9) | 4.2 (3.6) | +0.5 | Deterioration |
|  | Intervention | 132 | 4.3 (3.0) | 5.0 (3.3) | +0.7 | Deterioration |
| **HADS Depression/21**<br>Higher is inferior | Control | 132 | 3.9 (2.6) | 4.8 (3.7) | +0.9 | Deterioration |
|  | Intervention | 132 | 4.9 (2.7) | 5.3 (3.0) | +0.4 | Deterioration |
| **SHARE Frailty Index**<br>Higher is inferior | Control | 71 | 1.6 (1.6) | 1.7 (1.8) | +0.1 | Deterioration |
|  | Intervention | 72 | 1.7 (1.8) | 1.7 (1.8) | 0 | Deterioration |
| **Hand grip strength Right hand/kg**<br>Higher is inferior | Control | 78 | 24.9 (10.8) | 23.6 (9.6) | -1.3 | Deterioration |
|  | Intervention | 76 | 22.4 (8.3) | 20.9 (7.5) | -1.5 | Deterioration |
| **Carer Strain Index/13**<br>Higher is inferior | Control | 125 | 4.3 (3.3) | 4.7 (3.5) | 0.4 | Deterioration |
|  | Intervention | 134 | 4.7 (3.3) | 4.8 (3.5) | 0.2 | Deterioration |
| **Carer EQ-5D-5L Index/1.0**<br>Higher is superior | Control | 132 | 0.88 (0.17) | 0.86 (0.17) | -0.02 | Deterioration |
|  | Intervention | 140 | 0.85 (0.18) | 0.85 (0.16) | -0.01 | Deterioration |

Abbreviations: DAD Disability Assessment for Dementia, NEADL, Nottingham Extended Activities of Daily Living (NEADL), DEMQOL Dementia Quality of Life, LAPAQ Longitudinal Ageing Study Amsterdam Physical Activity Questionnaire, AES Apathy Evaluation Scale, EQ5D-3L EuroQol five dimensions questionnaire - three levels (EQ5D-3L), EQ5D-5L EuroQol five dimensions questionnaire – five levels, CSI Care Strain Index, MoCA Montreal Cognitive Assessment; HADS Hospital Anxiety and Depression Scale; SHARE Survey of Health Ageing and Retirement in Europe, MCID minimum clinically important difference

**Table 4:** Analysis of Covariance and standardised effect size estimates for intervention group (with missing data imputed)

| Measures | n | Adjusted mean difference (95% CI) * | Cohen's d effect Size (95%CI) * | p |
| --- | --- | --- | --- | --- |
| DAD Score | 365 | -1.3 (-5.2; +2.6) | -0.06 (-0.26; 0.15) | 0.51 |
| NEADL Score | 256 | +0.2 (-0.7; 1.1) | +0.05 (-0.20; 0.29) | 0.71 |
| LAPAQ Physical Activity Score | 365 | -167 (-445; 112) | -0.14 (-0.35; 0.06) | 0.25 |
| Accelerometer- Number of Steps in 7 days | 43 | -4030 (-11028; 2969) | -0.37 (-0.98; 0.23) | 0.25 |
| Berg Balance Scale | 145 | +1.8 (-0.7; 4.2) | +0.15 (-0.08; 0.57) | 0.15 |
| Timed Up and Go | 138 | -2.7 (-5.9; +0.5) | -0.29 (-0.62; +0.05) | 0.10 |
| Dual Task Timed Up and Go | 126 | -7.3 (1.8; 12.8) | -0.48 (0.12; 0.83) | 0.01 |
| DEMQOL | 365 | -3.8 (-6.8; -0.8) | -0.26 (-0.47; -0.06) | 0.01 |
| DEMQOL Proxy | 365 | +2.4 (-0.3; 5.1) | +0.18 (-0.03; 0.38) | 0.08 |
| DEMQOL-U (6 months) | 365 | +0.01 (-0.01; 0.04) | +0.11 (-0.1; 0.3) | 0.29 |
| EQ-5D-3L Index self-reported | 365 | +0.02 (-0.04; 0.07) | +0.08 (-0.12; 0.29) | 0.51 |
| EQ-5D-5L Index proxy | 365 | +0.03 (-0.01; 0.07) | +0.15 (-0.05; 0.36) | 0.16 |
| MoCA | 155 | -0.4 (-1.5; 0.8) | -0.11 (-0.42; 0.21) | 0.52 |
| Verbal Fluency - Correct Words | 155 | -0.5 (-1.6; 0.5) | -0.16 (-0.48; 0.15) | 0.32 |
| Apathy Evaluation Scale | 365 | -0.6 (-2.7; 1.4) | -0.07 (-0.27; 0.14) | 0.54 |
| Falls Efficacy Scale- International, short | 365 | +0.2 (-0.7; 1.0) | +0.05 (-0.15; 0.26) | 0.64 |
| HADS Anxiety | 275 | +0.4 (-0.3; 1.1) | +0.15 (-0.09; 0.38) | 0.23 |
| HADS Depression | 275 | -0.1 (-0.8; 0.6) | -0.03 (-0.27; 0.20) | 0.78 |
| SHARE Frailty Index | 149 | -0.05 (-0.52; 0.42) | -0.04 (-0.36; 0.29) | 0.56 |
| Hand Grip Strength /kg | 154 | -0.9 (-2.9; 1.1) | -0.15 (-0.47; 0.16) | 0.36 |
| Carer Strain Index | 365 | -0.01 (-0.63; 0.62) | -0.04 (-0.25; 0.16) | 0.69 |
| Carer EQ5D5L Index | 365 | +0.01 (-0.01; 0.04) | +0.09 (-0.12; 0.29) | 0.37 |

Note: Effect size Cohen's d interpretation: 0-0.2=no effect; 0.2-0.5=small; 0.5-0.8=moderate; >0.8=large
\*Positive values show an effect in favour of intervention group
Abbreviations: DAD Disability Assessment for Dementia, NEADL, Nottingham Extended Activities of Daily Living (NEADL), DEMQOL Dementia Quality of Life, LAPAQ Longitudinal Ageing Study Amsterdam Physical Activity Questionnaire, AES Apathy Evaluation Scale, EQ5D-3L EuroQol five dimensions questionnaire - three levels (EQ5D-3L), EQ5D-5L EuroQol five dimensions questionnaire – five levels, CSI Care Strain Index, MoCA Montreal Cognitive Assessment; HADS Hospital Anxiety and Depression Scale; SHARE Survey of Health Ageing and Retirement in Europe, CI Confidence Interval

The analyses of CANTAB cognitive measures were underpowered, but suggested statistically significant benefits for the PrAISED intervention, with a moderate effect size, on tests of multi-tasking (MTT, an executive function test assessing participants’ ability to manage conflicting information) and Spatial Span (SSP, a test of visuo-spatial working memory capacity; tables 5 and 6).

**Table 5:** CANTAB cognitive assessment results, unadjusted comparisons.

| Measure | Group | n | Baseline Mean (SD) | Follow-Up Mean (SD) | Difference | Interpretation |
| --- | --- | --- | --- | --- | --- | --- |
| **MTT RL median congruent/ms**<br>Higher is inferior | Control | 24 | 930 (168) | 953 (192) | 22 | Deterioration |
|  | Intervention | 26 | 904 (168) | 894 (163) | -9 | Improvement |
| **MTT RL median incongruent /ms**<br>Higher is inferior | Control | 24 | 1040 (193) | 1038 (193) | -1 | Improvement |
|  | Intervention | 26 | 1038 (173) | 993 (159) | -44 | Improvement |
| **MTT RL median switching blocks/ms**<br>Higher is inferior | Control | 24 | 1144 (246) | 1081 (192) | -63 | Improvement |
|  | Intervention | 26 | 1063 (240) | 1036 (183) | -27 | Improvement |
| **PAL memory score**<br>Higher is superior | Control | 27 | 3.5 (2.1) | 2.7 (2.1) | -0.8 | Deterioration |
|  | Intervention | 29 | 3.7 (3.4) | 3.0 (2.7) | -0.7 | Deterioration |
| **PAL number of patterns**<br>Higher is superior | Control | 27 | 4.9 (1.3) | 4.3 (1.3) | -0.6 | Deterioration |
|  | Intervention | 29 | 5.0 (1.5) | 4.4 (1.6) | -0.6 | Deterioration |
| **PAL total errors**<br>Higher is inferior | Control | 27 | 16.1 (7.4) | 13.9 (5.8) | -2.2 | Improvement |
|  | Intervention | 29 | 17.2 (7.3) | 15.5 (8.7) | -1.7 | Improvement |
| **SSP Lengths forward**<br>Higher is superior | Control | 28 | 4.1 (1.2) | 3.8 (1.3) | -0.3 | Deterioration |
|  | Intervention | 30 | 4.2 (1.2) | 4.5 (1.5) | 0.3 | Improvement |

MTT multi-tasking test (executive function); PAL paired associates learning (visual memory); SSP Spatial Span (visuospatial working memory); ES effect size, NS not statistically significant, CI confidence interval

**Table 6:** CANTAB cognitive assessment results, ANCOVA analysis.

| Measures | n | Adjusted mean difference (95% CI) | Effect Size (95%CI) | p-value | Interpretation (intervention) |
| --- | --- | --- | --- | --- | --- |
| MTT RL median congruent | 50 | -69 (-146; 7) | -0.58 (-1.14; +0.01) | 0.07 | Better, moderate ES, NS |
| MTT RL median incongruent | 50 | -79 (-159; 1) | -0.62 (-1.19; -0.05) | 0.05 | Better, moderate ES |
| MTT RL median switching blocks | 50 | -61 (-152; 30) | -0.43 (-0.99; + 0.14) | 0.19 | Better, small ES, NS |
| PAL memory score | 56 | 0.10 (-1.19; 1.39) | 0.05 (-0.48; +0.57) | 0.87 | Same |
| PAL number of patterns | 56 | 0.09 (-0.67; 0.84) | 0.07 (-0.46; +0.59) | 0.82 | Same |
| PAL total errors | 56 | 1.22 (-3.1; 5.55) | 0.16 (-0.37; +0.68) | 0.57 | Same |
| SSP lengths forward | 58 | 0.81 (0.14; 1.48) | 0.68 (+0.15; +1.2) | 0.02 | Better, moderate ES |

MTT multi-tasking test (executive function); PAL paired associates learning (visual memory); SSP Spatial Span (visuospatial working memory); ES effect size, NS not statistically significant, CI confidence interval

Multiple sensitivity analyses showed no differences in results, including (i) complete cases, (ii) those completing the intervention before the COVID-19 pandemic, (iii) using the interim data collected in the early weeks of the pandemic, (iv) correcting for survivor bias, by assigning a DAD score of zero to participants who died, (v) excluding those who terminated the intervention early due to the pandemic, (vi) excluding three participants who had MoCA scores at baseline above the upper limit.

4.4% of participants reported a confirmed COVID-19 infection. 82% engaged in social distancing for a median 116 days (IQR=37 to 210) and 47% COVID-19 self-isolated for a median 71 (IQR=22 to 139) days. Results were no different for those reporting, or not reporting, a COVID-19 infection.

### Falls

There were 796 falls in total, 375 in the intervention group and 421 in control group. 60% of participants in the intervention group had at least one fall in the trial compared with 57% participants in the control group, odds ratio, OR=1.1 (95% CI 0.71 to 1.8), p=0.6. There were 73 injurious falls in total, 38 in the intervention group, 35 in the control group. At least one injurious fall was reported by 15% of participants in the intervention and 16% in the control groups, OR=0.91 (0.51 to 1.6), p=0.8.

The falls efficacy outcome was falls in months 4-15. There were 279 falls recorded for participants who completed all the study calendars for these months, 79 in the intervention group, 200 in the control group. 59% of participants in the intervention group had at least one fall compared with 55% of participants in the control group, OR=1.2 (0.57 to 2.4), p=0.7. Falls incidence rate in the intervention group was 1.49 per person year compared with 2.47 per person years in the control group, Incidence Rate Ratio=0.78 (0.46 to 1.3), p=0.3, adjusted for site, co-resident carer and previous history of falls. A survival analysis showed median time to the first fall was 13 months for the intervention group, 12 months for the control group, adjusted Hazard Rate Ratio=0.85 (0.50 to 1.4), p = 0.5.

### Harms (adverse events)

One hundred and sixty-seven adverse events were recorded: 59 in control and 108 in the intervention groups, involving 68 participants: 27 (15%) from control and 61 (33%) from intervention groups. There were 91 serious adverse events: 29 in control and 62 in intervention, involving 60 participants: 22 (12%) from control and 38 (21%) from intervention groups. None was treatment related. There were 13 deaths: 4 (2.2%) in control and nine (4.9%) in the intervention groups (OR = 2.3, 95% CI 0.70 to 8.7, p = 0.3). There were 7 new care home placements; 2 (1.1%) from control and 5 (2.7%) from intervention group (OR = 2.4, 95% CI 0.49 to 19, p = 0.5). There were 75 hospital admissions: 27 in control and 48 in intervention groups, involving 53 participants: 22 (12%) from control and 31 (17%) from intervention groups (OR = 1.5, 95% CI 0.82 to 2.7, p = 0.2).

There were 228 falls recorded for participants who completed the first three months’ calendars, 96 falls in the intervention group and 132 falls in the control group. 32% of participants in the intervention group had at least one fall compared with 31% participants in the control group, OR = 1.1, 95% CI (0.64 to 1.8), p = 0.9.

## DISCUSSION

### Summary

The PrAISED intervention did not improve ADL, physical activity, quality of life or any other health status outcome, including balance and functional mobility, for people with mild dementia or MCI in the 12-month period after randomisation. There may have been a small reduction in rate of falling (22% relative risk, statistically uncertain), and improvement in some specific cognitive domains, in underpowered analyses, but these did not translate into functional gains. Delivery of the intervention was disrupted by COVID-19 pandemic restrictions.

### Strengths and limitations

This was a high-quality multi-centred RCT. We followed MRC guidance to develop and evaluate the intervention [83]. We established the feasibility and acceptability of intervention delivery and trial processes before starting the trial [22]. A process evaluation established reasonable participant adherence and fidelity of intervention delivery. Our attrition and missing data rates were acceptable. In qualitative interviews, undertaken as part of the process evaluation, the trial intervention was overwhelmingly well-received by participants, carers and provider staff [80].

The intervention was systematically designed, and refined over several years, including during the feasibly trial [21,22,54]. It was intended to be practical and relevant to participants. It comprised predominantly resistance (strength and balance) exercises, in a home setting, linking to daily activities, explicitly addressing risk of falls and other safety concerns and encouraging outdoor mobility. Intervention was individualised (tailored, personalised). Exercise was not a standard prescription but was seen as subserving activities that participants needed or wanted to do. Close attention was paid to motivation, the learning needs of people with dementia, and contextual factors, especially involvement of family or other carers. It was delivered by trained and experienced physiotherapists and occupational therapists, who made assessments and plans and supervised trained rehabilitation support workers.

The intervention was about as intensive as could be credibly delivered by a public health service. In designing the trial, the funder, NIHR, on behalf of the UK National Health Service, was concerned that the intervention was unfeasibly intensive, and requested the inclusion of a briefer and less expensive intervention in the feasibility study. The feasibility study demonstrated the need for prolonged supervision, however [22]. In our main trial, we emphasised tailoring of supervision to individual needs. Although a median of 31 therapy sessions over a year might have been insufficiently intensive to change outcomes (compared, for example, with the FINALEX study [19]) it was probably the maximum plausible dose in relation to NHS services and costs of delivery. We used ‘tapering’ of the intervention, with twice weekly visits in the first 3 months, reducing to monthly visits in the last 3 months to encourage independent undertaking of exercise. In the event, without direct supervision, this may simply have reduced adherence.

The patient population lacked diversity, being disproportionately well-educated, white men. The study enrolled people willing to agree to take part in research and perform prolonged exercise, who may have already been living healthy lifestyles and may have been the least likely to benefit.

The trial was disrupted by the COVID-19 pandemic, associated lockdown and social distancing restrictions. Clinical delivery teams were quick to move to remote delivery of the intervention and demonstrated great flexibility and innovation in doing so [71, 84], but this did not work for many participants, such as those with sensory impairments, lacking information technology hardware or internet connections, or a carer to help with telephone calls or videoconferencing. In this situation, progression of exercises was impossible to do safely, and access to community facilities diminished or ceased. Some follow-up interviews were conducted remotely, which might have affected data quality. Remote follow-up prevented us from collecting some secondary outcome measures. Sub-group analysis on participants followed-up prior to the COVID-19 pandemic did not suggest different results, however. Equally, it could be argued that the pandemic was challenging for all older people, our intervention could have mitigated this and shown exaggerated benefits [85,86].

We planned to objectively measure physical activities undertaken by participants in their own time using accelerometers but had to abandon this due to the pandemic [87]. We had no direct measure of participant independence, nor participant or carer satisfaction with the programme.

### Research in context

There have been numerous reviews of non-pharmacological interventions in dementia. The evidence that exercise and physical activity can improve ADLs for people with dementia is inconclusive. A Cochrane review found no high-quality evidence [16]. A further review concluded that exercise and physical activity reduced disability and falls, but that the quality of evidence was low [88]. A recent meta-analysis found no effect of exercise on ADLs [89]. Two reviews considered a range of interventions designed to maintain functional activity in dementia. Both identified heterogeneity between studies, mixed evidence of effectiveness, generally low quality of evidence, but a greater effect when interventions were tailored to participants’ interests and abilities, and delivered by registered therapists [90, 91]. The evidence for moderate to high intensity exercise preventing falls in cognitively intact older people is strong [92].

There have been a few adequately powered and high-quality individual trials. One trial of prolonged (12 months) and intensive (1 hour twice a week), physiotherapist-supervised, home exercise demonstrated a substantial reduction in rate of loss of ADL abilities and halved the rate of falling [19]. Two trials of exercise interventions for people with sarcopenia and frailty who were not cognitively impaired showed small but significant improvements in mobility disability incidence (20% risk reduction) and frailty markers with moderate intensity programmes [93-95]. Two 4-month trials of moderate to high intensity supervised group exercise for people with mild to moderate dementia showed no improvement in ADLs after six months [96,97]. Functionally orientated occupational therapy improved abilities and activity [60], but these findings were not replicated in two subsequent trials [98, 99]. The ‘Journeying Through Dementia’ trial of a bespoke, moderate-intensity, occupational therapy intervention was negative [100]. A trial of cognitive rehabilitation in mild to moderate dementia, focusing on functional activity, demonstrated that more goals were met in the intervention group, but no impact on health status measures such as ADL [101].

### Interpretation

Dementia is a progressive condition with no cure. In recent years, there has been increasing interest in preventing dementia [14]. Secular trends in incidence suggest that dementia risk is not immutable [6], but good evidence for the effectiveness of interventions to reduce dementia risk is lacking. Protective factors such as physical activity are likely to act over decades rather than months. ‘Secondary prevention’ (of progression once dementia is diagnosed) through lifestyle interventions appears to be ineffective. A reduction in rate of falling remains possible, and may be valuable, but did not impact on preservation of ADLs or quality of life. The point estimate of falls risk reduction in our study was in line with estimates from meta-analyses [92].

Current health policy emphasises living well after a dementia diagnosis, through a combination of healthcare, psycho-social and societal changes, and adaptation of services to meet the particular needs of people living with dementia [10]. We, and others, have demonstrated that maintaining abilities is not likely to be possible. This does not mean that intervention may not have benefit in the psycho-social domain, including affirming personhood, inclusion, occupation, relationships, or carer support. Aspects such as social engagement, concern, hope, goal-achievement, information-giving (on a range of dementia-related topics), and therapeutic relationship-building appear to have been greatly valued [80]. In palliative care and mental health, therapeutic relationships are valued in their own right; the exercise may have been a means to an end.

The absence of measurable health gain makes it hard to argue for routine provision of this intervention. However, our observations could inform the development of future models of support. There is a widespread perception of a ‘service gap’ for people after a dementia diagnosis and their carers. The healthcare background, knowledge and expertise of the therapists appears to have been relevant to delivering holistic and supportive intervention. What this means in terms of measurement and evaluation is yet to be defined, but our current paradigm may be missing something. Others have commented on the unsuitability of available outcome measures and the limitations of randomised controlled trials in evaluating interventions in this population [99, 100]. A Social Return on Investment analysis of our feasibility trial, a health economic methodology which attempts to identify, quantify and monetise a wide range of health, personal and social benefits from a public policy perspective, was strongly positive [102].

Some specific aspects of our trial may explain negative results. The DAD is recommended as the most appropriate ADL outcome for dementia trials but can be difficult to complete. It distinguishes between ‘initiation’ and ‘performance’ of activities, and privileges activities undertaken without prompting. Whilst reasonable on a normative basis, this may not adequately ascertain ‘supported performance’ rather than independence. We undertook training in dual-task-activities, as impairment in this is a risk factor for falling, it is trainable and improved abilities can carry over between activities [56-58]. Our main index of ability in dual tasking was the dual task TUG test, which deteriorated in the intervention group. The test involved getting up, walking three metres, turning and sitting down again, whilst counting backwards. The instructions were cognitively demanding. Researchers reported that participants who had received active therapy sometimes misunderstood the task, for example trying to walk backwards, and may have confused the test with therapeutic tasks they had practiced during the intervention. Similarly, the DEMQOL quality of life scale asks if the participant is ‘worried about’ things related to their dementia. The therapy programme may have increased participants’ awareness of their inabilities.

We can speculate whether control group participants, who get a diagnosis of dementia, and who are sufficiently able to volunteer for a research study, successfully adapt, drawing upon their existing resources and striking a balance that works for them in terms of activity and wellbeing. The objective of the trial was to introduce an intervention that did something different. Frequent involvement of health care professionals could disrupt normal adaption, draw attention to ill-health rather than well-being, encourage people to take greater risks than they would usually have done, and prompt greater involvement of health care services.

### Implications and future work

We add to accumulating evidence that interventions to delay cognitive or functional decline in early dementia or MCI are ineffective. So far, drug therapies, cognitive stimulation, exercise, and rehabilitation therapies have, at best, a small impact on functional activities and quality of life, and do not appear to change the course of the disease.

We need to think again about how we support people with dementia to live well with the condition. A more supportive approach to care may be required. Healthcare interventions should focus on solving practical problems and crises. We should emphasise helping the person with dementia to live well, despite their limitations, minimise intervention burden, maintain personhood, inclusion and occupation, provide psychological and emotional support, and support family and other carers.

Restoration of ‘independence’ in activities may be unrealistic, ‘adapted’ or ‘supported’ functioning (compensatory approaches) may be more achievable [103]. For example, a person may be assisted to cook or shop, in order that they remain active and included, rather than aiming for them to be able to do these tasks alone. We need outcome measures that reflect these. The value of therapeutic relationships may be underappreciated and may go beyond what might be expected from befriending, counselling or social prescribing. Exercise and physical activities should be promoted for enjoyment, occupation, inclusion and to enhance relationships.

To make a real impact on the course of dementia we are likely to need disease modifying drugs. Recent data on lecanemab provide proof of concept, but the ADL benefits over 18 months were very small (2 points on a 90-point scale) [104]. The failure of non-drug therapies justifies the continued search for new agents.

## Data Availability

All data produced in the present study are available upon reasonable request to the authors

## OTHER INFORMATION

### Trial Registration

ISRCTN15320670; prospectively registered 4-9-2018

### Protocol

https://trialsjournal.biomedcentral.com/articles/10.1186/s13063-019-3871-9

### Ethical Approval

This study was approved by the Bradford-Leeds Research Ethics Committee (REC number 18/YH/0059, IRAS project identification 236099) and research governance departments in each organisation.

### Data sharing

we are happy in principle to share our data. Please contact the corresponding author.

### Programme Steering Committee

S Iliffe (chair), L Allan, G Mountain, J Whitney, R Ogollah (statistician), M Lewis (statistician), P Riley (PPI), P Foster (PPI).

### Data Monitoring Committee

J Treml (chair), D Howell (statistician), A Bishop, E Mioshi.

## Authors contribution

- Conception, design, funding: RH, TM, SG, VvdW, PL, HS, KP, VHM, KV, RdN, MG, ZH, RTE, JG, KP, MO
- Intervention development, training and delivery support: VB, PL, TB, LH, AC, RT, JH, CDL, HS, LB, AL, MG, MD, TM, RH
- Literature searching: CDL, VB, VvdW, JH, FK, SG, RH
- Recruitment, data collection: RB, CB, JL, AL, ST, KJ, CD, CDL, AHS, TW
- Statistics, CTU: ZH, AB, RH
- Process evaluation: CDL, VvdW, KP, MG, MD, VB, LH, RH
- Health economic evaluation: RTE, VE, NH
- Study management: SG, VvdW, ROB, EA, MG, MD, KK, RV, TM, RH
- Interpretation: RH, CDL, VvdW, SG, FK, PL, JG, KP, VB, LH, AC, EA, RdN, TW, HS, MO, MG, MD
- Paper drafting: RH, SG, FK, CDL
- All authors approved the final paper

## Transparency statement

The lead author affirms that the manuscript is an honest, accurate, and transparent account of the study being reported; that no important aspects of the study have been omitted; and that any discrepancies from the study as originally planned have been explained.

