## Supplementary material for "Promoting Activity, Independence and Stability in early dementia and mild cognitive impairment (PrAISED): A randomised controlled trial"

**APPENDIX 1: Intervention description using TIDieR guideline**

<table>
<tr><td>1. Name of the intervention?</td><td>An intervention to Promote Activity, Independence, and Stability in Early Dementia and mild cognitive impairment (PrAISED)</td></tr>
<tr><td rowspan="3">2. Why do the intervention?</td><td>Intervention rationale:<br>People with dementia experience progressive deterioration in physical and mental abilities, due to increasing cognitive impairment, falls, acute and chronic co-morbidities, restricted opportunities, frailty and deconditioning. Exercise and rehabilitation therapies may improve abilities or reduce rate of decline, helping people to live well with dementia for longer.</td></tr>
<tr><td>What are the underpinning theories?<br>• Early intervention when participants can learn and develop new routines<br>• Exercise of the correct type, intensity and duration will increase capabilities, physical activity and reduce falls risk.<br>• Dual-task deficits can be reversed through training, reducing falls<br>• Gait re-education reduces falls risk<br>• Assessment, advice, adaptation and practice will improve performance of ADLs<br>• Assessment and advice on opportunities and risk management will improve performance of ADLs<br>• Environmental assessment for risk and safety promotes ADLs and reduce falls<br>• Implemented using dementia-specific motivational and behaviour-change theory will improve engagement and adherence</td></tr>
<tr><td>What is the aim of the intervention?<br>To promote activity and independence, and prevent falls, for older people with early dementia and mild cognitive impairment living in the community.</td></tr>
<tr><td rowspan="3">3. What materials are needed for the intervention?</td><td>Provider materials:<br>• Intervention manual<br>• Training programme</td></tr>
<tr><td>Participants materials:<br>• Paper copy of intervention plan and instructions in a home-file folder</td></tr>
<tr><td>Intervention equipment:<br>Variable-cuff-weights, household items such as a cup or glass, steps or stairs within the home, therapeutic balls and functional activities items</td></tr>
<tr><td rowspan="2">4. What procedures take place in the intervention?</td><td>Selection:<br>• Diagnosed dementia or MCI<br>• Montreal Cognitive Score of 13-25<br>• Living at home<br>• Ability and willingness to engage in a long-term exercise programme</td></tr>
<tr><td>Provider training:<br>• Group therapist training, 2-day initial (in person, digital during pandemic)<br>- Evidence base, 14 core principles, assessment, motivational strategies and goal setting<br>- Intervention content, ADLs, risk enablement, environmental access, physical activity, balance challenging, strength promotion, dual tasking, habit formation, tapering and fidelity.<br>• On-site mentoring<br>- Support within 2 months of commencement of intervention delivery or in response to specific site issues.<br>• Monthly peer-support group mentoring (remote, teleconference)</td></tr>
</table>

| | |
|---|---|
| | - Facilitated peer support, problem solving, sharing of best practice.<br>• Group Refresher Training Workshop, 1-day<br>- Intervention concepts including motivational strategies, dual tasking and ending of the intervention period<br>• Manual<br>- Intervention rationale, aims, assessments and content |
| | *Intervention session structure:*<br>• Initial session = therapist assessment, initial exercise tasks, clear information on programme, documentation<br>• Review session = feedback from the participant (programme, previous session, daily activities); functional activity (review goals and achievement); physical activity (prescribed exercises, physical activity outside the house); progression as appropriate; documentation of the session.<br>• Tailoring of structure through Frequency and Intensity Clinical Decision Tool |
| 5. **Who** is involved in the intervention? | *Intervention providers:*<br>• Registered (professionally qualified) Physiotherapists (PT) and Occupational Therapists (OT); support from unregistered Rehabilitation Support Workers (RSW)<br>• Experience working with either older people who fall or people with dementia<br>• Participated in the training programme before conducting intervention sessions |
| | *Participants:*<br>• Older adults (>65yrs) with mild dementia or mild cognitive impairment |
| 6. **How** is the intervention delivered? | • Face-to-face, one-to-one<br>• Intervention can be "paused" if circumstances demand (e.g. intercurrent illness or holiday) and participants placed on a review only pathway until ready to engage<br>• Carers or family members are invited to attend or participate in sessions, unless detrimental to the participant's engagement<br>• Motivational strategies are used to enhance adherence and uptake of the intervention<br>• Goal-setting is used to tailor the intervention to wishes/interests of participants<br>• Phone call by the clinician to supplement the intervention sessions |
| 7. **Where** is the intervention delivered? | • Participant's home<br>• Where functional or community activities require, intervention sessions may be conducted in the community, according to the participants goals and ability |
| 8. **When** is the intervention delivered? | • Some intervention sessions are supervised by OT, PT or RSW<br>• The intervention sessions facilitate independently undertaken physical activity for 150 minutes per week (i.e. 25 mins for 6 days of the week)<br>• Where possible, the day and time of regular supervised intervention sessions is maintained or agreed with the participant and carer |
| 9. How is the intervention **tailored**? | The programme is tailored according to clinical features, abilities, comorbidities, communication, interests and goals of the individual participant. |
| | *Tailoring occurs through:*<br>• Content (physical and functional activities, motivational strategies)<br>• Delivery (day, time, number of sessions, who delivers sessions)<br>• Goal-setting (functional activities, interests, prior experiences, aims, when the goals are set, terminology used) |

| | |
|---|---|
| | *Progression is achieved through:*<br>• Maintaining an achievable challenge for the individual<br>• Increasing physical difficulty (number of repetitions or time completing each exercise; resistance for strength exercises; reducing base of support for balance exercises)<br>• Increasing cognitive difficulty (difficulty of the dual-task component; complexity of functional task; reducing number of prompts or adaptions)<br>• Reducing support (frequency of supervised sessions)<br>• Content/tailoring is established during the initial sessions of the programme and monitored by the qualified therapists and/or RSW |

Hoffmann TC, Glasziou PP, Boutron I, Milne R, Perera R, Moher D, Altman DG, Barbour V, Macdonald H, Johnston M, Lamb SE, Dixon-Woods M, McCulloch P, Wyatt JC, Chan AW, Michie S. Better reporting of interventions: template for intervention description and replication (TIDieR) checklist and guide. BMJ 2014; 348:g1687. DOI: 10.1136/bmj.g1687.

**APPENDIX 2. Guidance distributed to the PrAISED therapists on changes to be made to the intervention as a result of the COVID-19 restrictions (issued 18-3-2020)**

**Immediate plan**

The NIHR have stated that their funded studies should stop all non-essential face-to-face contact. The PrAISED intervention is not considered essential care and therefore we must stop all face-to-face contact with our participants.

However, because we have a duty of care to our patients considering many of them will be following the government's advice to reduce all social contact, we have devised a contingency plan to continue with the PrAISED intervention.

**Intervention Group Participants**

Therapy teams should contact all participants currently in the trial, or their carers if more appropriate, to explain the change in practice as below.

**On-going Intervention Group Participants**

Visits to participants should be replaced with **telephone coaching** as per their normal schedule, in terms of frequency. For example, if you are seeing someone weekly, this should be continued until they reach the time to reduce to fortnightly. This is the example frequency schedule set out in the intervention manual, however, continue to adapt this as appropriate in the same way you have been doing.

- Month 1-2: bi-weekly
- Month 3-6: weekly
- Month 6-9: fortnightly
- Month 9-12: monthly

The length of the phone call may be much shorter depending on what is discussed.

The content of the phone call should be guided by the telephone coaching instructions below.

Some participants won't be suitable for telephone calls. If the participant is unable to engage with telephone coaching the carer should be contacted to determine if they may be able to use the telephone coaching to support the participant. If the telephone coaching is of no benefit to either the participant or the carer, then a courtesy telephone call should be given each month to keep in touch with the carer or participant as appropriate.

Final sessions should be carried out via the telephone as appropriate; these should be followed up with an end-of-therapy letter and any follow up material being provided using the post or email if appropriate.

**New Intervention Group Participants**

Intervention group participants seen by the research team but not yet seen by therapy team, or who are in the assessment phase of the intervention, should be informed that they are not going to receive the PrAISED intervention until the current restrictions are lifted.

**Control Group Participants**

If you have completed the first control visit you can carry out up to two follow up visits by telephone as per the guidance below. If the first control visit has not yet been completed, please inform the participant that they are not going to receive the PrAISED intervention until the current restrictions are lifted.

**Therapy Visit Log**

Continue to complete the therapy visit log, via the hyperlink, for all telephone calls. Please put telephone coaching in the comments box.

**Medium-Term Plan**

It is expected that PrAISED therapy staff at each site will deliver the immediate plan outlined above.

However, as the situation changes a medium-term plan (outlined below) may come into action.

If sites cannot deliver the telephone coaching sessions due to therapy staffing difficulties, the university staff may have capacity to be able to support. The PI from each site must contact the University as soon as possible if this happens. For university staff to be able to do the telephone coaching sessions effectively, we will need to know:

- the participant's details (e.g., contact telephone number for them and the carer/informant)
- a synopsis of the previous intervention session and what they are currently working on

As each site is using different participant documentation systems, the PIs should liaise with Sarah Goldberg or Rebecca O'Brien, to form a contingency plan on how this will happen and how information is to be transferred and stored.

**Telephone Coaching Instructions**

Before making the telephone call make sure you have looked at NHS England current advice for the client group you are dealing with, as this is likely to change on a regular basis (https://www.nhs.uk/conditions/coronavirus-covid-19/). Participants may have concerns about their current situation that need answering before the participant will engage in coaching.

- Explain who you are and why you're calling.
- Ask how they are and discuss any immediate concerns (they may need signposting as appropriate).
- Review their current activity and exercise plan.
- Review what they are currently doing during their day.
- Be aware that for many participants all their activities may have stopped.
- Form a plan of what they can do within the **current** restrictions. For example, currently people are advised it is ok to walk outside as long as they stay 2m away from other people.
- Help them to make a daily plan of activities. For example, doing exercises more frequently, or if they are no longer walking outside can they walk in the garden or up and down the stairs to get some cardiovascular exercise.

- Advise against sitting for long periods of time. For example, use a timer to remind yourself to get up or get up during advert breaks in television programmes.
- If the person is able to and wants to, they could put you on speaker phone while you go through their exercise programme with them.  Only do this if they have the capacity to do this with their telephone.  This could also be done with their carer or family member or named informant.
- Be aware people may be feeling quite worried and/or low in mood.  You may need to discuss the benefits of, and encourage them to continue to carry out daily activities or routines, such as getting dressed, or taking meals on time.
- Participants may raise safeguarding issues such as identifying they are low on medication and there is no one to help them with this. This will need to addressed using the usual safeguarding procedures.
- If participants are complaining of COVID 19 symptoms they should be encouraged to follow the current advice from NHS direct or to phone 111.

It is expected these telephone coaching guidelines will evolve as PrAISED therapists start conducting these sessions.  Guidance can come from outside sources, e.g., RCOT have recently shared this online https://www.rcot.co.uk/staying-well-when-social-distancing.  It is important that we share good practice and suggestions and will discuss these guidelines during our PrAISED Therapist Teleconferences.

**APPENDIX 3: Data collection on fidelity, adherence, reach, dose and adaptations**

| | **Training** | | **Intervention** | |
|---|---|---|---|---|
| | Delivery (PrAISED team) | Attendance (Therapists) | Delivery (Therapists) | Adherence (Participants) |
| **Reach** | Number of sites receiving training: Recorded by study team* | Number of therapists attending training and number of therapists completing training tasks: Attendance record of core training and questionnaire filled in by individual therapists on core training days* | Number of visits: Visit logs compiled by individual therapists after each visit and provided to study team, who checked for any errors in entering data* | Number of participants who completed the programme, their baseline characteristics (including sex, age, ethnicity, marital status) and cognitive score (MoCA); and number of participants withdrawn from study: Recorded by study team* |
| **Dose** | Days / hours of training per site. Attendance record of core training, teleconferences, on-site monitoring visits and therapist conference were compiled by the study team* | Days / hours of attendance per therapist: Attendance record at core training, teleconferences, on-site monitoring visits and therapist conference were compiled by the study team* | Days / hours of visit: Visit logs compiled by individual therapists, collected by local sites and provided to study team who checked for any errors in entering data* | Minutes per week recorded on calendar: Participants were asked to record on monthly calendars provided by the study team their daily minutes of PrAISED-related activities and return the calendars to the study team* |
| **Fidelity /adher-ence** | Delivery of training as planned: Training sessions were recorded and checked against Standard Operating Procedure* | Attendance at training as planned: Attendance record of core training, teleconferences, on-site monitoring visits and therapist conference were compiled by the study team* | Delivery of intervention against PrAISED principles: Process evaluation team recorded a convenience sample of therapists (n = 14) delivering one therapy session each in the participant's home at month six of the therapists' involvement. Videos were assessed against the 14 principles the therapists received training on during the core training | Minutes per week recorded on calendar: Participants were asked to record on monthly calendars provided by the study team their daily minutes of PrAISED-related activities and return the calendars to the study team* |
| **Adapt-ations** | Any adaptations made when providing training: Recorded by study team* | | Goals for participants and any adaptations made to tailor the programme: Visit log compiled by individual therapists recorded goals and changes* | Adaptations that participants made to the original programme: Information gathered through qualitative interviews of process evaluation |

* Data collected during the RCT
